# Aging-related Changes in Bimanual Coordination As a Screening Tool for Healthy Aging

**DOI:** 10.1101/2025.01.06.25320094

**Authors:** Yusuke Shizuka, Shin Murata, Akio Goda, Shun Sawai, Shoya Fujikawa, Ryosuke Yamamoto, Takayuki Maru, Kotaro Nakagawa, Hideki Nakano

## Abstract

The steady increase in the global older adult population highlights critical challenges, including the development of preventive strategies to extend healthy life expectancy and support independence in activities of daily living. Although there is an aging-related reduction in manual dexterity, the difference in bimanual coordination performance between young and older adults remains unclear. We aimed to elucidate the characteristics of bimanual coordination among young, young-old, and old-old adults participants. The participants performed in-phase (tapping the thumb and index finger together as fast as possible) and anti-phase (alternating movement between the left and right fingers) bimanual coordination tasks, and intergroup comparison of the task parameters as performed. The number and frequency of taps significantly decreased sequentially in young, young-old, and old-old adults, whereas the average tap interval significantly increased in this order (*p* < 0.05). There was no significant difference between the young-old and old-old groups in the average local maximum distance (*p* > 0.05). These findings indicate that bimanual coordination task performance varies depending on specific parameters. Thus, the number, average interval, and frequency of taps are potential indicators of aging-related changes in bimanual coordination.

## 1. Introduction

In recent years, the proportion of older adults in the global population has steadily increased. The United Nations World Social Report 2023 indicates that the number of people aged 65 years and above would more than double, from 761 million, in 2021, to 1.6 billion, by 2050. Likewise, the population aged 80 years and above is expected to grow rapidly. In 2021, one in ten people in the global population was 65 years or older, and by 2050, this could increase to one in six people [1]. Compared with younger adults, older adults tend to experience a decline in physical function. Aging-related deterioration in motor function significantly contributes to increased caregiver burden, higher hospitalization rates, and increasing healthcare costs [2, 3]. Consequently, interest in promoting healthy aging has been growing. Healthy aging is defined as the process of developing and maintaining functional abilities that enable wellbeing in older age [4]. Furthermore, the relationship between healthy aging and physical activity has been underscored with regard to potential contributions to pain alleviation and prevention of falls, osteoporosis, sarcopenia, and cognitive impairments [5]. Thus, healthy aging is essential to maintain the ability to perform activities of daily living (ADL). Therefore, in the global population wherein the number of older adults is increasing annually, it is crucial to implement preventive measures to promote healthy aging, extend healthy life expectancy, and support the ADL independence of older adults.

The upper limbs are frequently used in daily life, and older adults perform more bimanual, than unilateral, tasks [6]. Therefore, maintaining and improving finger function are crucial for older adults to lead healthy lives. Incel et al. found that grip and pinch strengths in older adults are associated with activity limitations and quality of life [7]. Moreover, maximal grip strength and motor performance involving coordinated bimanual control decrease with age, which suggests that older adults may find it challenging to perform daily activities that require bimanual movements, such as holding a bottle while simultaneously unscrewing its cap [8]. Aging-related cognitive decline is a relevant risk factor [9]. Rattanawan reported that older adults with cognitive impairment tended to experience a decline in ADL performance owing to reduced manual dexterity and impaired bimanual coordination [10]. Therefore, finger function plays a vital role in ADL performance, with a particularly strong correlation between bimanual tasks and daily activity levels in older adults. Thus, to promote healthy aging and support independence in older adults, it is essential to appropriately assess finger function during bimanual movements and undertake preventive interventions to prevent aging-related decline.

A meta-analysis investigating aging-related changes in bimanual movement among older adults revealed a decline in accuracy, increased variability, and prolonged execution times for bimanual coordination [11]. Another meta-analysis investigating the characteristics of bimanual coordination in older adults found that motor performance deterioration was more pronounced in asymmetrical, rather than in symmetrical, bimanual tasks [12]. Thus, it has been clarified that bimanual coordination performance in older adults declines in comparison to that in young adults. Furthermore, both young-old adults and old-old adults exhibit reduced bimanual performance compared to younger adults [13, 14]. Asymmetrical bimanual coordination performance is lower in young adults than in older adults, with performance decline particularly pronounced in tasks that require higher movement speeds [13]. The accuracy of both symmetrical and asymmetrical bimanual force control is lower in old-old adults, compared to young adults [14]. Therefore, bimanual coordination performance, which plays a crucial role in daily activities, decreases in both young-old and old-old adults as compared to that in young adults. Typically, old-old adults experience more pronounced decline in physical and cognitive functions than young adults [15-17]. Therefore, although aging-related changes in bimanual coordination may occur in young-old and old-old adults, the differences in bimanual coordination performance between young-old and old-old adults remain unclear.

We aimed to clarify the age-specific characteristics of bimanual coordination in young, young-old, and old-old adults. The research hypothesis was that bimanual coordination performance would be lower in older adults than in young adults, and that among older adults, the decline in performance would be more pronounced in old-old adults than in young-old adults. If the age-specific characteristics of bimanual coordination can be clarified, these tasks can be applied as screening tools for health management and promotion among older adults. Moreover, early detection and intervention for the decline in ADL among older adults could contribute to promoting healthy aging and extending their health span.

## 2. Materials and Methods

### 2. 1. Materials

Participants included 107 healthy younger adults and 364 healthy older adults. The exclusion criteria were as follows: (i) older adults under 65 years; (ii) older adults with suspected cognitive impairment based on a Mini-Mental State Examination (MMSE) score of 23 or less [18]; (iii) participants with musculoskeletal, central nervous system, or mental disorders that may have affected the study results; (iv) left-handed participants; and (v) participants whose maximum amplitude of distance was 300 mm or more in bimanual coordination measurements [19], (vi) Participants who could not complete all measurements correctly. After excluding the participants who met the exclusion criteria, 97 healthy younger adults and 324 healthy older adults were included in the analysis (Figure 1). Additionally, participants were categorized into three groups: young adults (18–22 years), young-old adults (65–74 years), and old-old adults (≥75 years) [20].

**Figure 1.**
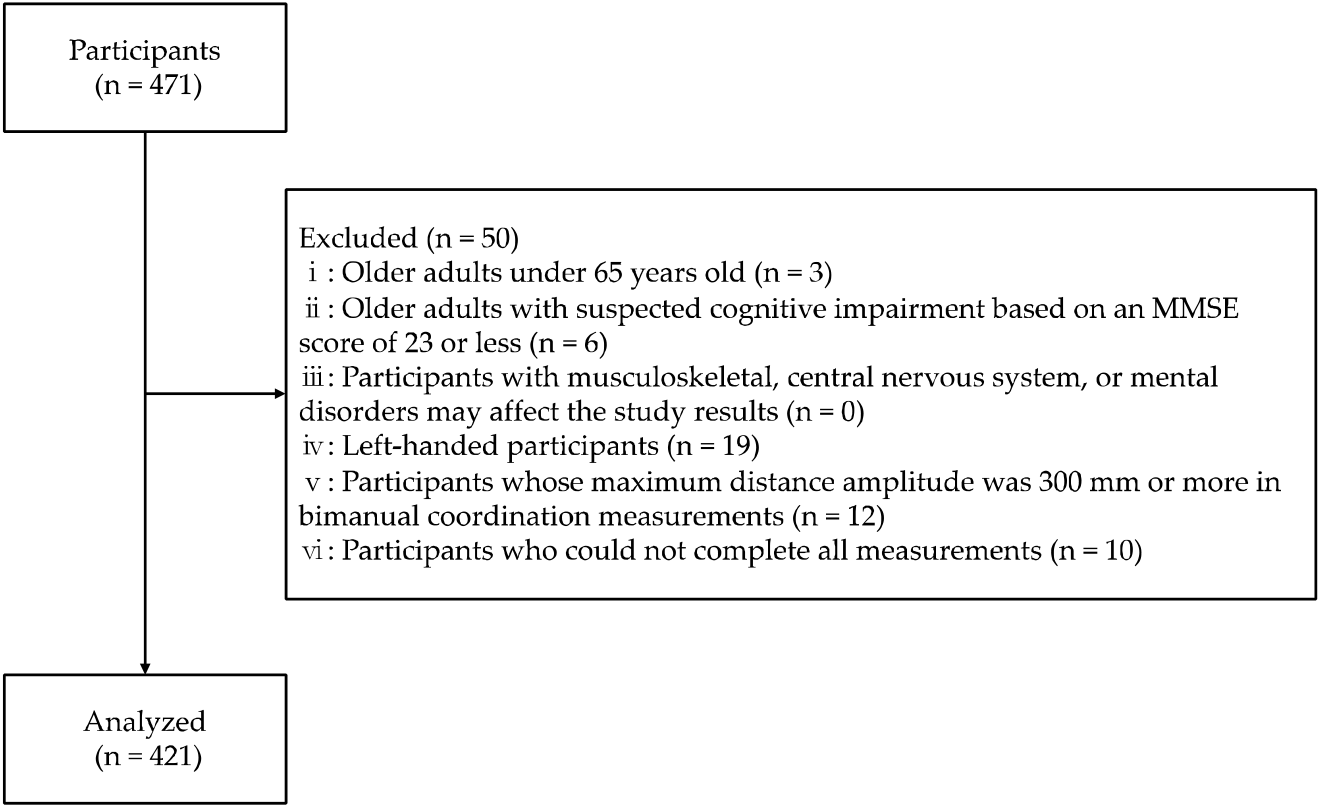
Flowchart of participation criteria. MMSE: Mini-Mental State Examination.

### 2. 2. Ethics statement

This study was conducted in accordance with the principles of Declaration of Helsinki and approved by the Research Ethics Committee of Kyoto Tachibana University (approval number: 24-52, approval date: November 12, 2024). Informed consent was obtained from all the participants. The study was registered in the UMIN Clinical Trials Registry (UMIN000056499).

### 2. 3. Methods

All the participants performed a bimanual coordination task involving opposition movements of the thumb and index finger [21] in two tasks: the in-phase task, where tapping movements of the thumb and index finger were performed simultaneously and as quickly as possible with both hands; and the anti-phase task, where tapping movements were alternated between the left and right hands [22] (Figure 2), wherein participants sat on chairs with backrests and placed their forearms on the platform. During each task, the forearms were positioned in a neutral rotation, with the third, fourth, and fifth fingers slightly flexed, and measurements were taken with the eyes closed (Figure 3). We instructed the participants to “perform as fast as possible and in the same rhythm.” The tasks were performed in sequence, starting with the in-phase task and followed by the anti-phase task. Each task was conducted for 15 s, and a 15-s pre-practice session was provided before each task.

**Figure 2.**
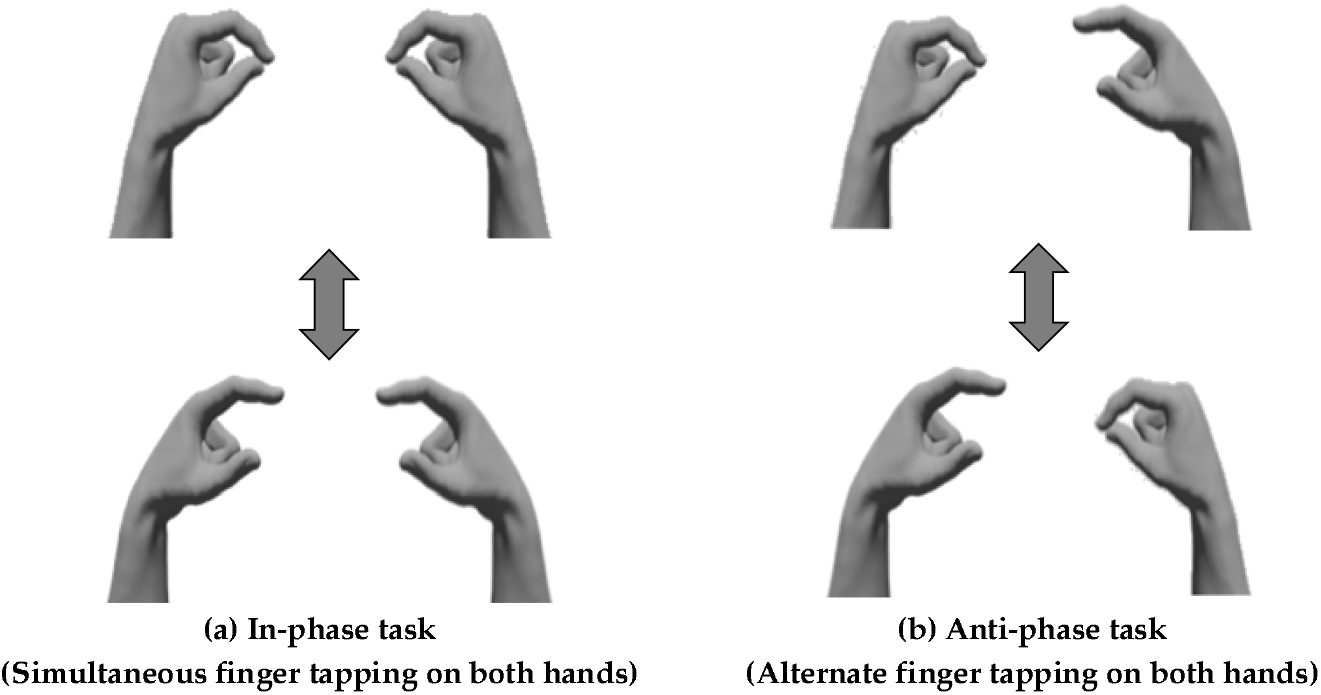
Bimanual coordination task. (a) In-phase task: Participants performed tapping movements of the thumb and index finger simultaneously on both sides. (b) Anti-phase task: Participants performed tapping movements of the thumb and index finger alternately on both sides.

**Figure 3.**
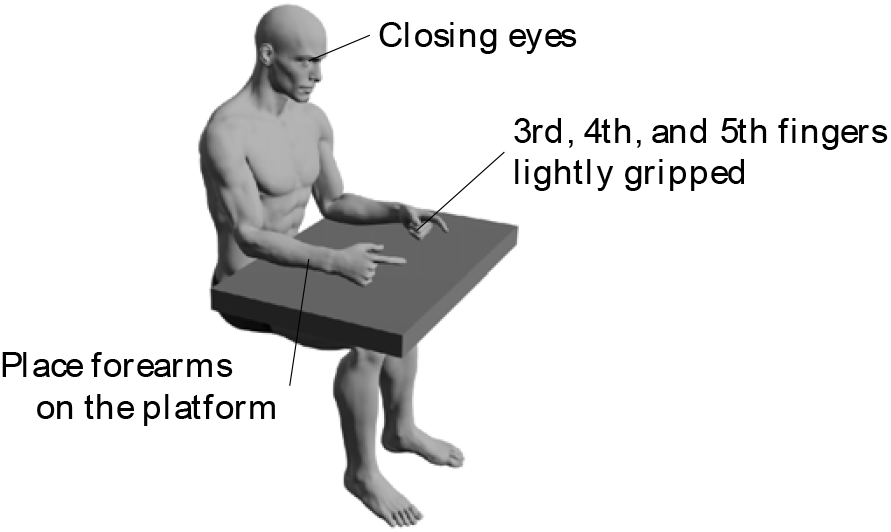
Measurement position of the bimanual coordination task. Participants sat on chairs with backrests and placed their forearms on the platform. During each task, the forearms were positioned in a neutral rotation, with the third, fourth, and fifth fingers slightly flexed, and measurements were taken with the participants sitting with eyes closed.

Bimanual coordination performance was measured using a magnetic sensor finger-tapping device (UB-2, Maxell Ltd. Tokyo, Japan) [21]. Magnetic sensors were attached to the dorsal side of the participant’s thumb and index finger with a rubber band and the distance from the strength of the magnetic field generated between the two fingers. This device was highly reproducible and reliable across periods, devices, and measurement examiners [23]. In this study, the participants were instructed to open their fingers to a width of 40 mm to minimize amplitude variation, and both pre-practice and measurements were conducted [24]. The features of bimanual coordination were obtained from the recorded data [22] (Table 1), which yielded four parameters for evaluating the distance and movement amplitude of the thumb and index fingers during the task. Tap interval-related parameters yielded four parameters for evaluating the average speed of movement and variability of tapping. The phase difference-related parameters yielded one parameter to evaluate the timing discrepancy of tapping between the hands. For each parameter, larger values of the total traveling distance, average local maximum distance, number of taps, and frequency of taps indicated better bimanual coordination performance. Conversely, smaller values for the standard deviation of the local maximum distance, slope of the approximate line of local maximum points, average of tap intervals, standard deviation of the inter-tap interval, and standard deviation of the phase difference indicate better performance for bimanual coordination. We instructed the participants to open their fingers to a width of 40 mm. Therefore, the closer the mean of the average of the local maximum distance is to 40 mm, the better the performance of bimanual coordination.

**Table 1.**
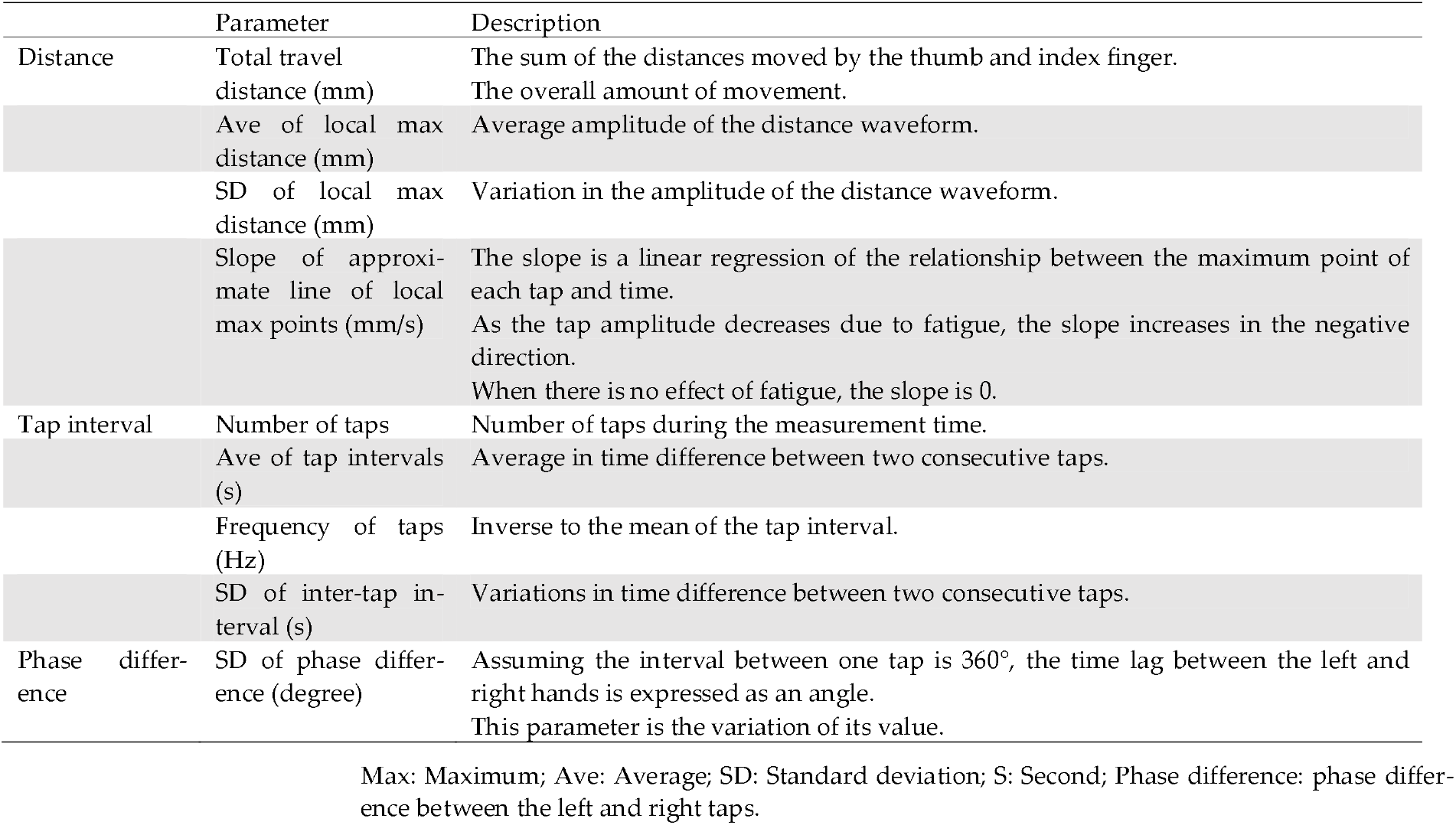
Characteristics of the bimanual coordination task.

|  | Parameter | Description |
| --- | --- | --- |
| Distance | Total travel distance (mm) | The sum of the distances moved by the thumb and index finger.<br>The overall amount of movement. |
|  | Ave of local max distance (mm) | Average amplitude of the distance waveform. |
|  | SD of local max distance (mm) | Variation in the amplitude of the distance waveform. |
|  | Slope of approximate line of local max points (mm/s) | The slope is a linear regression of the relationship between the maximum point of each tap and time.<br>As the tap amplitude decreases due to fatigue, the slope increases in the negative direction.<br>When there is no effect of fatigue, the slope is 0. |
| Tap interval | Number of taps | Number of taps during the measurement time. |
|  | Ave of tap intervals (s) | Average in time difference between two consecutive taps. |
|  | Frequency of taps (Hz) | Inverse to the mean of the tap interval. |
|  | SD of inter-tap interval (s) | Variations in time difference between two consecutive taps. |
| Phase difference | SD of phase difference (degree) | Assuming the interval between one tap is 360°, the time lag between the left and right hands is expressed as an angle.<br>This parameter is the variation of its value. |

Max: Maximum; Ave: Average; SD: Standard deviation; S: Second; Phase difference: phase difference between the left and right taps.

### 2. 4. Statistical Analysis

First, a chi-square test was conducted to compare the male/female ratios among the young, young-old, and old-old adults. Next, a three-way ANOVA with a mixed design was conducted to compare the parameters related to the distance and tap interval, considering the factors of the hand (left, right), task (in-phase task, anti-phase task), and group (young, young-old, and old-old adults). A two-way mixed-design ANOVA was used to compare the parameters related to the phase difference, considering task (in-phase task, anti-phase task) and group (young, young-old, and old-old adults) factors. Bonferroni post-hoc tests were performed for parameters with significant interactions or main effects using ANOVA. SPSS version 29.0 (IBM Corp., Armonk, NY, USA) was used for the statistical analysis, with the significance level set at 5%.

## 3. Results

### 3. 1. Basic Information of Participants

The 421 participants were divided into groups based on age: 97 young adults (male: 25, female: 72, age: 20.73 ± 1.43 years), 102 young-old adults (male: 17, female: 85, age: 70.77 ± 2.70 years), and 222 old-old adults (male: 52, female: 170, age: 80.71 ± 4.38 years). The results of the chi-square test showed no significant differences between male and female ratios among the young, young-old, and old-old adult groups (*p* > 0.05).

### 3. 2. Comparison of the Distance Parameters

The statistical analysis revealed no significant three-way interaction (hand × task × group) for any distance parameter (*p* > 0.05; Table 2). The total traveling distance showed significant interactions between the hand × group and task × group factors (*p* < 0.05), and the total traveling distance had a significant effect on the task factor (*p* < 0.05). Post-hoc test results indicated that the total traveling distance in the old-old adult group was significantly lower for the left hand than for the right hand (*p* < 0.05). Moreover, the total traveling distance was significantly greater during the in-phase task than during the anti-phase task for the young, young-old, and old-old adult groups (*p* < 0.05). The total traveling distance in the anti-phase task was significantly lower in the old-old adult group than in the young and young-old adult groups (*p* < 0.05).

**Table 2.**
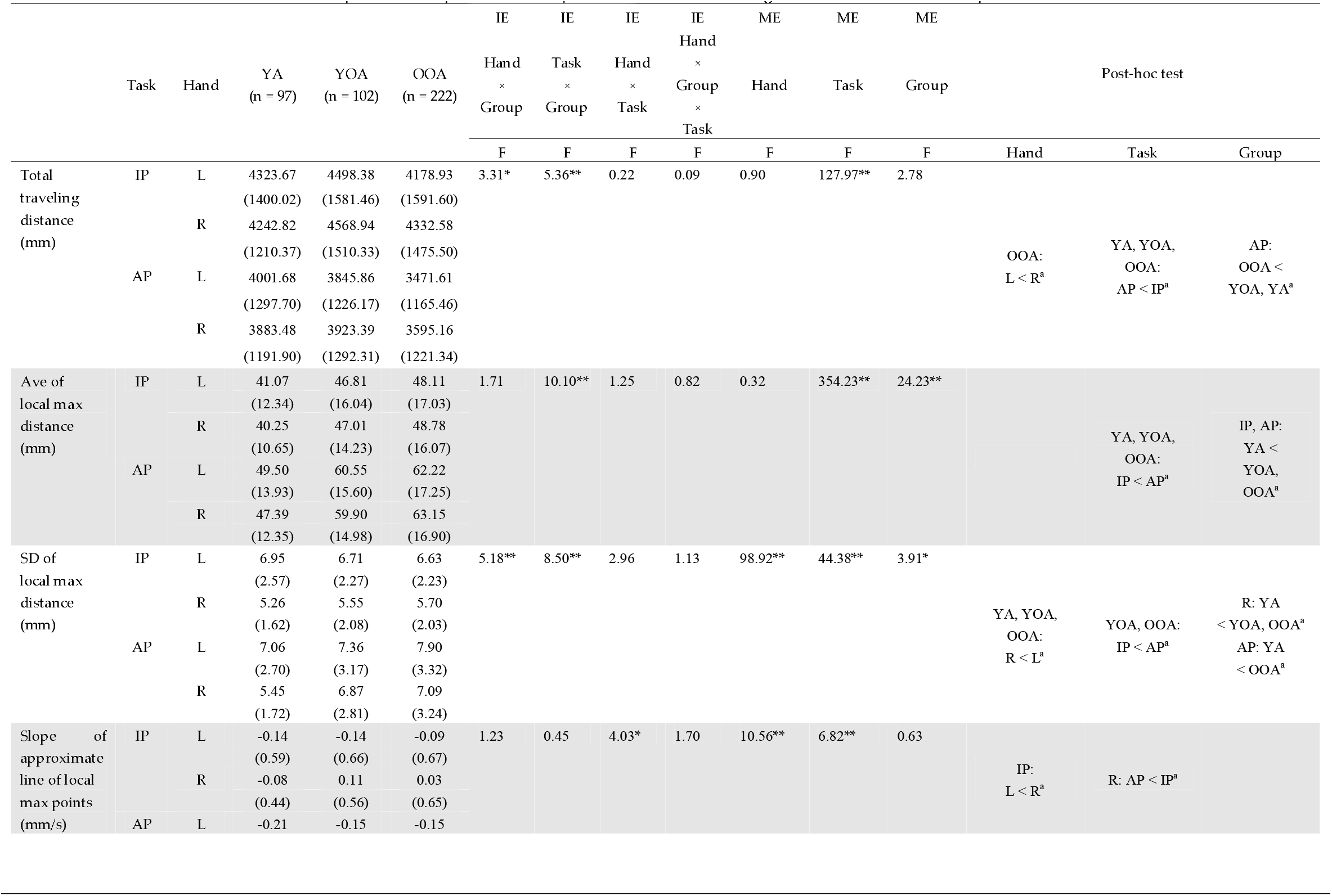
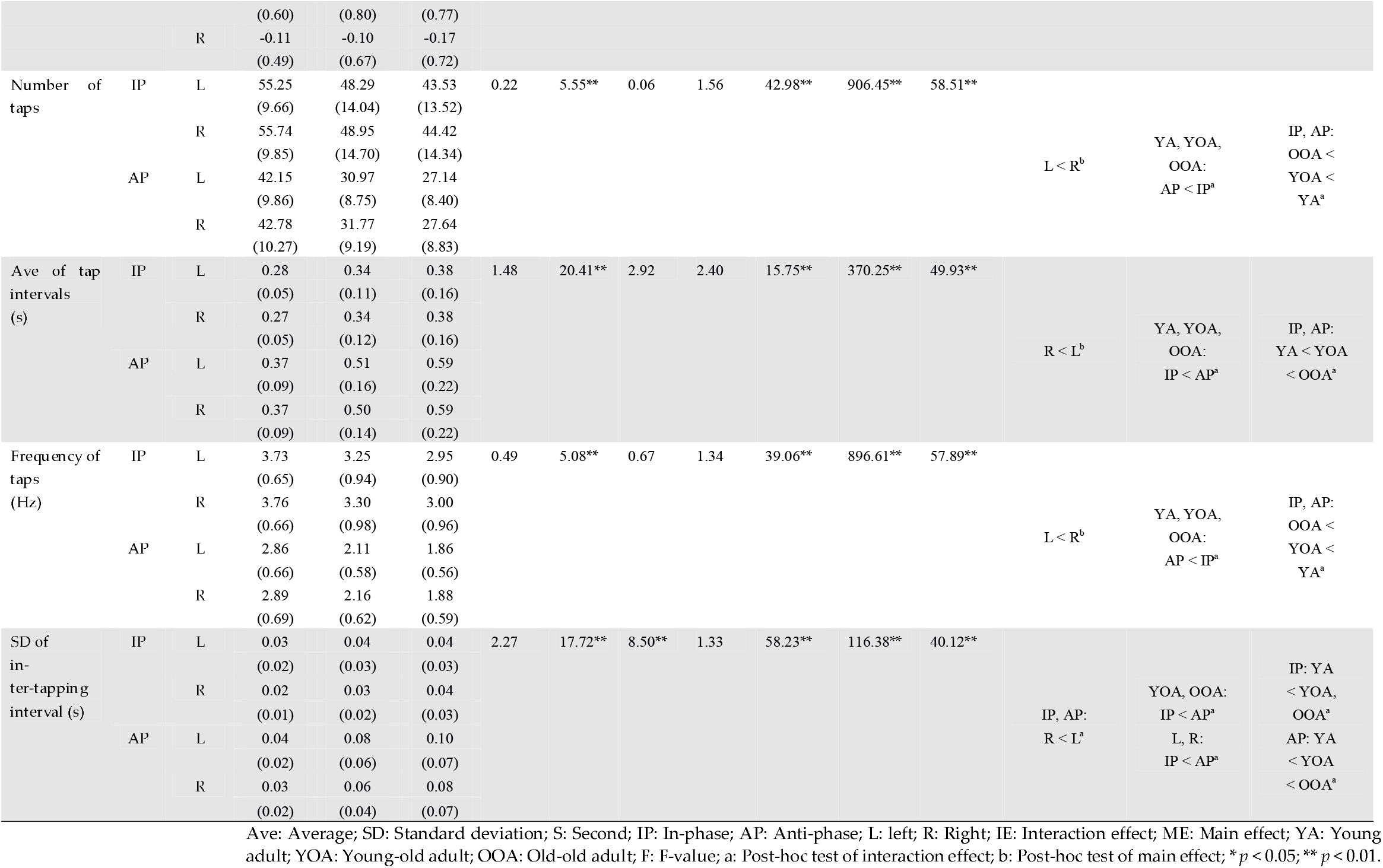
Comparison of In-phase and Anti-phase tasks and Left and Right hands at Distance and Tap interval.

The average local maximum distance showed a significant interaction for the task × group factor (*p* < 0.05). The average local maximum distance showed significant main effects for both task and group factors (*p* < 0.05). The results of post-hoc tests indicated that the average local maximum distance was significantly greater during the anti-phase task than during the in-phase task for the young, young-old, and old-old adult groups (*p* < 0.05). Furthermore, the average local maximum distance was significantly higher in the young-old and old-old groups than in the young adult group during the in-phase and anti-phase tasks (*p* < 0.05).

The standard deviation of the local maximum distance showed significant interactions for both hand × group and task × group factors (*p* < 0.05). Moreover, the standard deviation of the local maximum distance showed significant main effects for hand, task, and group factors (*p* < 0.05). The results of post-hoc tests indicated that the standard deviation of the local maximum distance was significantly higher for the left hand than for the right hand in the young, young-old, and old-old adult groups (*p* < 0.05), and the standard deviations were significantly greater in the anti-phase task than in the in-phase task (*p* < 0.05). Furthermore, the standard deviation of the local maximum distance for the right hand was significantly higher in the young-old and old-old adult groups than in the young adult group. The standard deviation of the local maximum distance in the anti-phase task was significantly higher in the old-old adult group than in the young adult group (*p* < 0.05).

The slope of the approximate line of the local maximum points showed a significant interaction for the hand × task factor (*p* < 0.05). Additionally, the slope of the approximate line of the local maximum points showed significant main effects for both hand and task factors (*p* < 0.05). Post-hoc tests indicated that the slope of the approximate line of local maximum points in the in-phase task for the left hand was significantly lower than that for the right hand (*p* < 0.05). Furthermore, the slope of the approximate line of the local maximum points for the right hand was significantly lower in the anti-phase task than that in the in-phase task (*p* < 0.05).

### 3. 3. Comparison of the Tap Interval Parameters

Statistical analysis revealed no significant three-way interaction (hand × task × group) for any of the tap-interval parameters (*p* > 0.05; Table 2).

The number of taps showed a significant interaction effect between task and group factors (*p* < 0.05). The number of taps showed significant main effects for the hand, task, and group factors (*p* < 0.05). Post-hoc analysis indicated that the number of taps was significantly lower for the left hand than for the right hand (*p* < 0.05). Furthermore, the number of taps in the young, young-old, and old-old adult groups was significantly lower in the anti-phase task than in the in-phase task (*p* < 0.05). Finally, the number of taps in both the in-phase and anti-phase tasks decreased significantly in the following order: young adults, young-old adults, and old-old adults (*p* < 0.05).

The average tap intervals showed a significant interaction effect between the task and group factors (*p* < 0.05). Additionally, the average tap intervals showed significant main effects for hand, task, and group factors (*p* < 0.05). Post-hoc test results indicated that the average tap interval was significantly longer for the left hand than for the right hand (*p* < 0.05). Moreover, the average tap intervals in the young, young-old, and old-old adult groups were significantly longer in the anti-phase task than in the in-phase task (*p* < 0.05). Finally, the average tap intervals in both the in-phase and anti-phase tasks significantly increased in the following order: young adults, young-old adults, and old-old adults (*p* < 0.05).

The frequency of taps showed a significant interaction effect between task and group factors (*p* < 0.05). Additionally, the frequency of taps showed significant main effects for hand, task, and group factors (*p* < 0.05). Post-hoc analysis test results indicated that the frequency of taping was significantly lower in the left hand than in the right hand (*p* < 0.05). Furthermore, the frequency of taps in the young, young-old, and old-old adult groups was significantly lower in the anti-phase task than in the in-phase task (*p* < 0.05). Lastly, the frequency of taps in both the in-phase and anti-phase tasks decreased significantly in the following order: young adults, young-old adults, and old-old adults (*p* < 0.05).

The standard deviation of the intertap intervals showed significant interaction effects for the task × group and hand × task factors (*p* < 0.05). Additionally, the standard deviation of the inter-tap intervals showed significant main effects for hand, task, and group factors (*p* < 0.05). Post-hoc test results indicated that the standard deviation of inter-tap intervals was significantly higher for the left hand than for the right hand in both the in-phase and anti-phase tasks (*p* < 0.05). Moreover, the standard deviation of the inter-tap intervals in the young-old and old-old groups was significantly higher in the anti-phase task than in the in-phase task for both hands (*p* < 0.05). Finally, the standard deviation of inter-tap intervals in the in-phase task was higher in the young-old adult group and the old-old adult group than in the young adult group, whereas it significantly increased in the order of young adults, young-old adults, and old-old adults in the anti-phase task (*p* < 0.05).

### 3. 4. Comparison of the Phase Difference Parameters

The standard deviation of the phase difference showed a significant interaction effect for the task × group (*p* < 0.05). Additionally, the standard deviation of the phase difference showed significant main effects for both task and group factors (*p* < 0.05). The Post-hoc test results indicated that the standard deviation of the phase difference for the young-old and old-old adult groups was significantly higher in the anti-phase task than in the in-phase task (*p* < 0.05). The standard deviation of the phase difference in the anti-phase task increased significantly in the following order: young adults, young-old adults, and old-old adults (*p* < 0.05; Table 3).

**Table 3.**
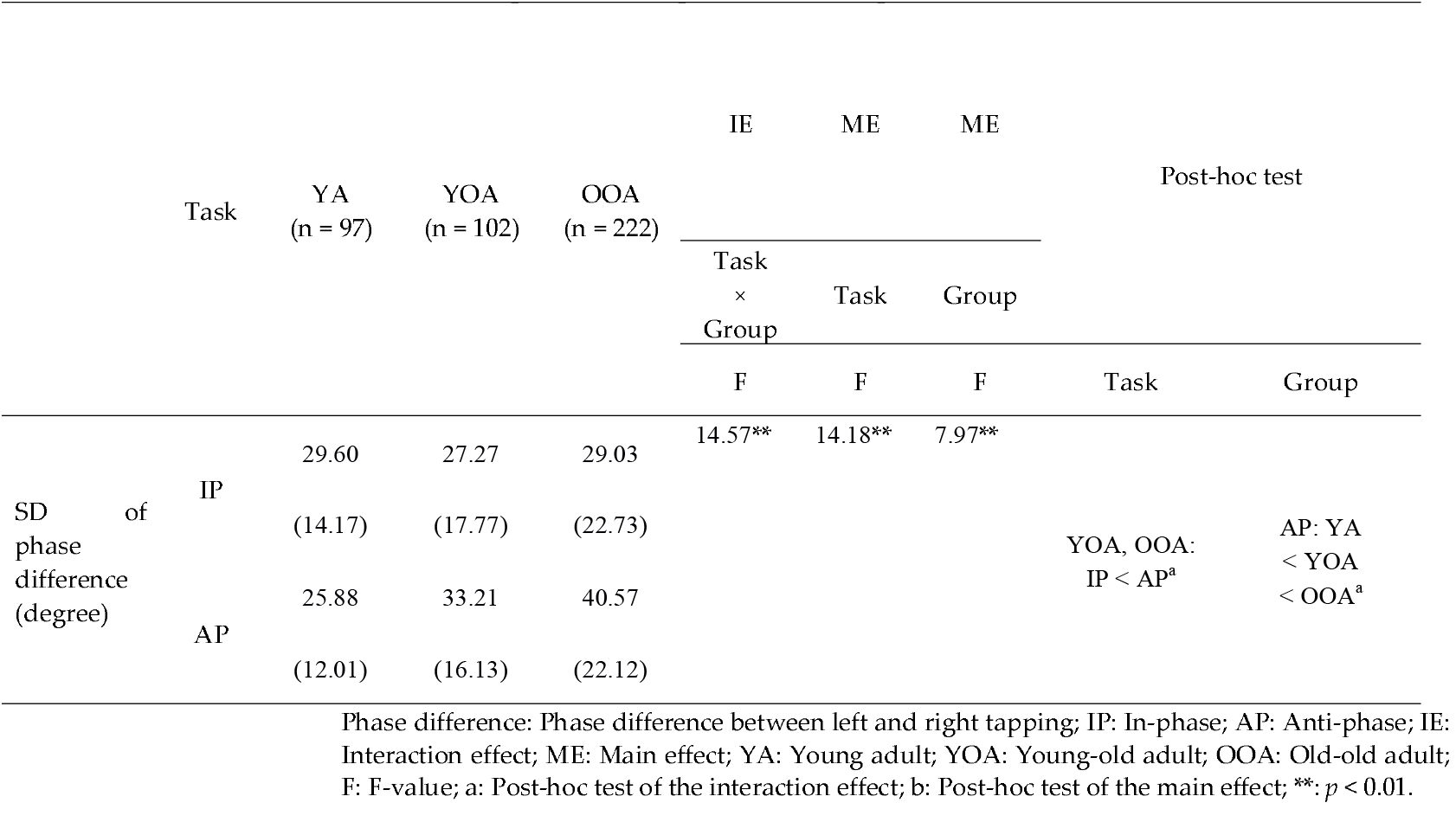
Comparison of In-phase and Anti-phase tasks at Phase difference.

## 4. Discussion

This study aimed to clarify age-related changes in bimanual coordination by comparing the performance of young, young-old, and old-old adults on bimanual coordination tasks. The results showed that the performance on the bimanual coordination task was lower in the older adult group than in the young adult group. In particular, the number and frequency of taps decreased, while the average tap interval increased in the following order: young adults, young-old adults, and old-old adults. On the other hand, the average local maximum distance and the standard deviation of local maximum distance increased more in the older adult group than in the young adult group, but remained consistent between the young-old and old-old adult groups. Furthermore, the performance of the anti-phase task was lower than that of the in-phase task, and the left-hand performance was lower than that of the right hand. These results suggest that the number of taps, mean tap interval, and frequency of taps are potential tools for assessing age-related changes in bimanual coordination.

### 4.1. Comparison of Young Adults, Young-Old Adults, Old-Old Adults

In this study, the performance of bimanual coordination tasks was compared among young, young-old, and old-old adult groups. The parameters were categorized into those that changed with aging among the young adult, young-old adult, and old-old adult groups, and those that showed differences between the young adult and older adult groups but no difference between the young-old adult and old-old adult groups.

The number and frequency of taps decreased in the order of young adult, young-old adult, and old-old adult groups during both in-phase and anti-phase tasks. Conversely, the average tap intervals increased in the order of young adult, young-old adult, and old-old adult groups during both the in-phase and anti-phase tasks. This indicates that aging is associated with a reduction in the number of taps and an increase in the time required for a single-tapping action. A study comparing bimanual performance between young and young-old adults reported that the young-old adult group exhibited a reduction in the number of finger taps and an extension of movement time, leading to a decline in task performance compared to the young adult group [13]. Furthermore, in a bimanual force regulation task, aging was found to increase performance errors, leading to a decline in the ability to control coordinated forces with both hands in young, young-old, and old-old adult group [25]. In summary, aging appears to impair bimanual coordination and reduce both movement quantity and accuracy. Consequently, it is possible that, in this study, the number of taps decreased and the time required for a single-tapping action increased in the order of young adults, young-old adults, and old-old adults. Additionally, interhemispheric interactions via the corpus callosum play a crucial role in bimanual coordination [26], and age-related structural and functional changes in the corpus callosum have been reported to impair bimanual performance [27, 28]. These findings suggest that the performance of bimanual coordination tasks declined sequentially from the young adult group to the young-old adult group to the old-old adult group, potentially because of age-related impairments in the interaction between the left and right cerebral hemispheres.

The standard deviation of the inter-tap intervals increased during the anti-phase task in the following order: young adult, young-old adult, and old-old adult. This indicates that the variability in the time required for a single tapping action during an anti-phase task increases with age. Previous studies examining age-related changes in bimanual force control during in- and anti-phase tasks have reported greater variability in the time taken to exert the specified force among older adults compared with young adults, with a particularly pronounced decline in performance during anti-phase tasks compared with in-phase tasks [14]. Furthermore, age-related impairments in neuromodulation in the brain can lead to random activation of neurons, resulting in increased intraindividual performance variability [29]. Therefore, in addition to changes in motor performance due to task difficulty, increased variability in motor performance resulting from age-related impairments in neural regulation within the brain led to a progressive increase in the standard deviation of the inter-tap interval from the young adult group to the young-old adult group and from the young-old adult group to the old-old adult group. Similarly, the standard deviation of phase differences during the anti-phase task increased in the order of the young adult, young-old adult, and old-old adult groups. This indicated that the timing discrepancy between the tapping movements of the right and left hands increased with age. The accuracy of bimanual coordination declines in older adults compared with young adults [11]. Additionally, a study on nerve conduction velocity in the peripheral nervous system, spanning ages 20 to 103 years, revealed a linear decrease in the average conduction velocity with age in both males and females [30]. In this study, it is possible that the peripheral nerve conduction velocity declined with age. Consequently, the timing discrepancy in tapping movements between the right and left hands likely increased in the order of the young, early elderly, and late elderly groups. Furthermore, Rhythmic bimanual coordination stability is influenced by neural crosstalk, movement amplitude, and conduction time delays [31]. Therefore, peripheral nerve conduction velocity may have declined with age in this study. In summary, the age-related decline in peripheral nerve conduction velocity may have impaired rhythmic bimanual coordination, resulting in a progressive increase in the standard deviation of the phase difference from the young adult group to the young-old adult group, and from the young-old adult group to the old-old adult group. In this study, differences between the groups in the standard deviations of the inter-tap intervals and phase differences were observed only during the anti-phase task. The supplementary motor area plays a crucial role in the bilateral motor control during bimanual coordination. Previous studies have shown that in healthy young adults, activity in the supplementary motor area is more prominent during anti-phase movements than during in-phase movements. However, motor facilitation connectivity within cortical motor networks, including the supplementary motor area, decreases with age [32, 33]. These findings suggest that decreased activity of the supplementary motor area involved in anti-phase movements due to aging may have caused the observed differences in the variability of single-tap movements and interhand coordination, specifically in the anti-phase task, among young adults, young adults, and old-old adults.

The average local maximum distances significantly increased in the young-old and old-old adult groups compared to the young adult group during both the in-phase and anti-phase tasks. This indicates that the opening width between the thumb and index finger increased in the older adult group compared to the young adult group, regardless of task difficulty. Motor control in bimanual coordination declines with age, and accuracy decreases in older adults compared to young adults [11]. Additionally, working memory, which is the ability to temporarily store and actively process information to achieve specific goals, has been reported to decline with age [34]. In the bimanual coordination task used in this study, the participants were instructed to maintain a finger separation width of 40 mm between the thumb and index finger and were required to perform the tasks while preserving this interval. Therefore, in this study, the decline in working memory due to aging may have prevented participants from maintaining a consistent distance between the thumb and index finger, leading to an increase in the average local maximum distance in the older adult group compared with the young adult group.

The standard deviation of the local maximum distances increased in the right hand for the young-old and old-old adult groups compared to the young adult group. This indicated that the variability in the opening width of the fingers was greater in the older adult group than in the young adult group for the right hand. Bimanual coordination variability has been reported to be higher in older adults than in young adults [11]. Moreover, proprioception in the fingers declines with age. Older adults show longer proprioceptive reaction times and reduced position sense than younger adults [35]. It is possible that the age-related decline in position sense made it difficult for older adults to consistently maintain the opening width of their fingers. Thus, the variability in finger-opening width during the bimanual coordination task may have been greater in the older adult group than in the younger adult group. Additionally, studies on unilateral and bilateral upper limb targeting tasks have shown that the task accuracy is generally higher for the right hand than for the left hand [36]. Therefore, the variability in the opening width of the fingers during bimanual coordination tasks may have been reduced in the dominant right hand compared to the non-dominant left hand. These findings suggest that the standard deviation of the local maximum distances was larger in the older adult group than in the young adult group for the right hand.

The standard deviation of inter-tap intervals increased in the in-phase task for the young-old and old-old adult groups compared with the young adult group. This indicates that the variability in the time required for a single-tapping action was greater in the older adult group than in the young adult group during the in-phase task. Previous studies have reported that motor performance in bimanual coordination declines significantly in older adults compared to young adults [13, 14]. Additionally, older adults exhibit a trade-off between movement frequency and motor control accuracy. They tended to compensate for the reduced accuracy by decreasing the frequency of bimanual coordination movements [13]. In this study, tapping frequency, which represents the frequency of single-tapping actions, decreased in the following order: young adults, young-old adults, and old-old adults. However, the standard deviation of inter-tap intervals, representing variability in the time required for a single-tapping action, increased in the older adult group compared with that in the young adult group. Moreover, in this study, the slope of the approximate line of the local maximum points, which reflects fatigue, decreased more during the anti-phase task than during the in-phase task. This suggests that fatigue develops more easily in the in-phase task than in the antiphase task. Additionally, the number of taps performed by the older adult group was higher in the in-phase than in the anti-phase task. Thus, it is possible that in older adults, the increased number of taps during the in-phase task compared to the anti-phase task led to greater fatigue, which in turn resulted in increased variability in the time required for a single-tapping action. These findings suggest that the standard deviation of the intertap interval may have been higher in older adults than in young adults during in-phase tasks.

In summary, this study demonstrated that the performance on bimanual coordination tasks declines in older adults compared to young adults. The average of local max distances and standard deviation of local maximum distances, which reflect the distance of the bimanual coordination task, were greater in the older adult group than in the young adult group. Furthermore, among older adults, the number and frequency of taps decreased in the following order: young, young-old, and old-old. Conversely, the average tap intervals increased in the following order: young adults, young-old adults, and old-old adults. These findings suggest that the number of taps, average tap intervals, and the frequency of taps, which reflect the speed of bimanual coordination, could serve as useful indicators for assessing age-related changes in bimanual coordination performance among young, young-old, and old-old adults.

### 4.2. Comparison of the In-phase and the Anti-phase tasks

In this study, the total traveling distance, number of taps, and frequency of taps decreased during the anti-phase task compared to the in-phase task among young, young-old, and old-old adults. Conversely, the average local maximum distance and the average tap intervals increased during the anti-phase task compared with the in-phase task among young adults, young-old adults, and old-old adults. This indicates that, regardless of the participants’ age, the amount of hand movement and the number of taps decreased during the anti-phase task compared to the in-phase task, whereas the time required for a single tap and the range of hand opening increased during the anti-phase task compared to the in-phase task. In the in-phase task, the tapping movements of the thumb and index finger were performed with both hands, and in the anti-phase task, the tapping movements were alternated between the left and right hands. The anti-phase task requires different movements of the left and right hands, making it more challenging than the in-phase task. Consequently, bimanual coordination performance is expected to decline in the anti-phase task compared to the in-phase task. It has been shown that bimanual motor impairments increase during asymmetrical bimanual tasks like the anti-phase task compared to symmetrical tasks like the in-phase task, and these impairments are more pronounced in older adults than in young adults [12]. Research on behavioral principles in inter-limb and hand-foot coordination has reported that during anti-phase movements, interference from one hand’s movement affects the other hand’s movement, resulting in spatial and temporal constraints. As the frequency of anti-phase movements increases, there is a tendency to shift toward in-phase movements for symmetry [37]. In other words, in the in-phase task, the movements of one hand likely facilitated the movements of the other hand, enhancing performance in the bimanual coordination task. In contrast, in the anti-phase task, the movements of one hand may interfere with those of the other, leading to reduced performance in the bimanual coordination task. Indeed, in this study, the number and frequency of taps increased in the less demanding in-phase task and decreased in the more demanding anti-phase task. Thus, the performance of bimanual coordination tasks appears to depend on task difficulty. This suggests that both the young adult and older adult groups in this study likely experienced a decline in performance during the anti-phase task compared to the in-phase task. Furthermore, it has also been reported that in young adults, the greater the micro-structural integrity in the midbrain regions, the better the motor performance in tasks requiring interactions between the cerebral hemispheres [38]. Additionally, compared with young adults, older adults tend to have smaller anterior fibers of the corpus callosum and poorer performance on anti-phase bimanual coordination tasks [39]. These findings suggest that in addition to task difficulty, the reduced amount of movement and number of taps observed during the anti-phase task in this study may be influenced by the structural integrity of the corpus callosum fibers in young and older adults, particularly in tasks requiring alternating bimanual coordination.

Furthermore, in this study, the standard deviation of the inter-tap interval increased during the anti-phase task compared with the in-phase task for both the left and right hands. This indicates that the variability in the time required for a single-tapping movement was greater in the anti-phase task than in the in-phase task. It has been reported that the performance of bimanual coordination task—such as accuracy, variability, and execution time—decreases during asymmetrical tasks compared to symmetrical tasks [21, 39, 40]. Asymmetrical tasks are generally considered more challenging than symmetrical tasks. The standard deviation of the inter-tap interval, which is one of the parameters in this study, serves as an indicator of variability in bimanual coordination tasks. An in-phase task corresponds to a symmetrical task, whereas an anti-phase task corresponds to an asymmetrical task. These findings suggest that temporal variability in the time required for a single-tapping motion increased in the anti-phase task compared to the in-phase task because of the influence of task difficulty.

In addition, the standard deviations of the local maximum distance, inter-tap interval, and phase difference increased during the anti-phase task compared to the in-phase task in both the young-old and old-old adult groups. This indicates that, in the older adult group, the variability in the range of hand opening, time required for a single-tapping movement, and timing discrepancies between the hands were greater during the anti-phase task than during the in-phase task. It has been reported that the variability in bimanual coordination increases with age, with older adults exhibiting more pronounced differences between in-phase and anti-phase tasks than younger adults. While no differences in the performance of bimanual coordination tasks were observed between the two tasks in young adults, older adults tended to experience reduced performance on the bimanual coordination task during the anti-phase task compared to the in-phase task [39]. This suggests that the reduction in corpus callosum size and the decline in its microstructural integrity with aging may have led to an increase in the standard deviation of local maximum distance, standard deviation of inter-tap interval, and standard deviation of phase difference in the older adult group during the anti-phase task compared with the in-phase task, but not in the young adult group. This indicates that older adults’ performance on bimanual coordination tasks tends to be more dependent on task difficulty than younger adults. It is also more prone to decline during high-difficulty antiphase tasks, which require greater interhemispheric interactions.

On the other hand, the slope of the approximate line of the local maximum points decreased in the right hand during the anti-phase task compared to the in-phase task. This indicated that the anti-phase task was less influenced by fatigue than the in-phase task. Indeed, in this study, the number and frequency of taps decreased, while the average tap interval increased during the anti-phase task compared to the in-phase task. In other words, the in-phase task involved shorter tap intervals and a higher number of taps than the anti-phase task, which may have made it more prone to fatigue. A study comparing maximal grip strength and endurance between the dominant and nondominant hands reported that while the dominant hand exhibited greater absolute strength, it was more susceptible to early fatigue [41]. Furthermore, it has been noted that the muscle fiber composition in the fingers of the dominant hand includes a significantly higher proportion of Type II fibers, which excel in rapid force generation, compared to Type I fibers, which are better suited for endurance [42]. These findings suggest that the reduced endurance of the dominant right hand resulted in a decreased slope of the approximate line of local maximum points during the anti-phase task compared to the in-phase task.

In summary, this study revealed that the performance on bimanual coordination tasks declined during the anti-phase task compared to the in-phase task. The number of bimanual coordination tasks decreased during the anti-phase task among all age groups, including young, young-old, and old-old adults. In contrast, spatial and temporal variability in bimanual coordination tasks was greater in the anti-phase task than in the in-phase task only in the older adult group. These findings suggest that the performance of bimanual coordination tasks may decrease during the anti-phase task compared to the in-phase task among young, young-old, and old-old adults. Furthermore, spatial and temporal variability in bimanual coordination tasks tended to increase more prominently in young-old and old-old adults than in young adults.

### 4.3. Comparison of the Left hand and Right hand

In this study, the total traveling distance, number of taps, and frequency of taps were lower for the left hand than for the right hand. However, the standard deviation of the local maximum distance, average of tap intervals, and standard deviation of the intertap interval were higher for the left hand than for the right hand. This indicates that the left hand has less finger movement compared to the right hand and shows greater variability in the distance between the thumb and index finger, as well as in the time required for each tapping motion compared to the right hand. Repetitive movement tasks may have been suggested to induce changes in activity within the motor cortex, a phenomenon known as use-dependent plasticity [43]. Research on handgrip strength and finger dexterity between the dominant and non-dominant hands has reported significantly stronger grip strength and higher finger dexterity in the dominant hand than in the non-dominant hand [44]. Notably, all the participants in this study were right-handed. Therefore, it is possible that use-dependent plasticity resulted in lower performance in the left hand than in the right hand across many parameters in the bimanual coordination task. Furthermore, the total traveling distance was lower in the left hand than in the right hand in the old-old adult group. This indicates that finger movement in the old-old adult group was lower in the left hand than in the right hand. It has been reported that finger usage of the dominant hand increases with age compared with that of the non-dominant hand [45]. Therefore, age-related increases in finger usage might have caused a left-right difference in finger movement in the old-old adult group, the oldest age group. These findings suggest that bimanual coordination performance may be lower in the older adult group than in the young adult group and lower in the non-dominant left hand than in the dominant right hand.

Next, the standard deviation of the inter-tap interval was higher for the left hand than for the right hand for both the in-phase and anti-phase tasks. This indicated that the variability in the time required for a single tapping movement was greater in the left hand, regardless of the task type. In general, right-handed individuals exhibit higher manual dexterity in their dominant hand than in their non-dominant [45]. Therefore, the standard deviation of the inter-tap interval, which represents temporal variability, increased in the non-dominant left hand compared with the dominant right hand owing to use-dependent plasticity.

Furthermore, the standard deviation of the local maximum distance was higher for the left hand than for the right hand across all age groups, including young, young-old, and old-old adults. This indicated that the variability in the distance required for a single tapping movement was greater in the left hand than in the right hand, regardless of age. A previous study that used tasks that required drawing in-phase and anti-phase circles with both hands reported that movement variability was lower in the dominant hand than in the non-dominant hand [46]. Therefore, the standard deviation of the local maximum distance, which represents spatial variability, is thought to have increased in the non-dominant left hand compared to the dominant right hand.

The slope of the approximate line of the local maximum points during the in-phase task was lower for the left than for the right hand. This indicated that the left hand experienced less fatigue than the right hand during the in-phase task. The in-phase task was considered less challenging than the anti-phase task, and the increased movement during the task may have led to fatigue. A previous study reported that during tasks requiring simultaneous maximal force exertion with both hands, the dominant hand demonstrated a faster decline in force than the non-dominant hand [47]. In this study, the number and frequency of taps were lower in the left hand than in the right hand and were also lower during the anti-phase task than during the in-phase task. Similarly, the average tap intervals were higher in the left hand than in the right hand, and higher during the anti-phase task than during the in-phase task. These results suggest that the right hand in the in-phase task may have been the most fatigued hand because it moved the most during the task.

In summary, it was revealed that the performance of bimanual coordination task was lower in the non-dominant left hand compared to the dominant right hand. Notably, unlike the young and young-old adult groups, the old-old adult group showed significantly less movement in the left hand than in the right hand during the bimanual coordination task. These findings suggest that the left-right difference in the amount of movement in bimanual coordination tasks may increase with age.

### 4.4. Limitations

This study had some limitations. First, age-related changes in bimanual coordination were examined only at the behavioral level, leaving the neural mechanisms in the brain unclear. Second, this study focused solely on age-related changes in bimanual coordination using tapping tasks, and its relationship with other physical and psycho-physiological functions remains uncertain. In recent years, bimanual coordination tasks have been increasingly used as screening tests for cognitive impairment, and it has been suggested that they may also be associated with other physical and psychological functions [48]. Therefore, future studies should aim to clarify the neural mechanisms involved in bimanual coordination and their relationship with other physical and psychophysiological functions, using techniques such as transcranial magnetic stimulation and electroencephalography.

## 5. Conclusions

We clarified age-related changes in bimanual coordination by comparing the performance of bimanual coordination tasks among young, young-old, and old-old adults. The performance of bimanual coordination task was lower in the non-dominant left hand than in the dominant right hand during the more challenging anti-phase task compared to the less challenging in-phase task. Furthermore, the features of the bimanual coordination task revealed parameters that showed minimal variations between young-old and old-old adults as well as among parameters that exhibited significant aging-related changes. Specifically, the number and frequency of taps decreased, whereas the average tap intervals increased in the following order: young, young-old, and old-old adults. These findings suggest that the number of taps, the average of tap intervals, and the frequency of taps may be useful as tools for assessing aging-related changes in bimanual coordination.

## Author Contributions

Conceptualization, S.M. and H.N.; methodology, S.M. and H.N.; formal analysis, Y.S.; investigation, Y.S., S.M., A.G., S.S., S.F., R.Y. and H.N.; data curation, Y.S.; writing—original draft preparation, Y.S.; writing—review and editing, Y.S., S.M., A.G., S.S., S.F., R.Y.,T.M., K.N. and H.N; visualization, Y.S.; supervision, S.M. and H.N; project administration, S.M. and H.N; funding acquisition, S.M., T.M., K.N. and H.N. All authors have read and agreed to the published version of the manuscript.

## Institutional Review Board Statement

This study was conducted in accordance with the Declaration of Helsinki and approved by the Ethics Committee of Kyoto Tachibana University (approval number: 24-52; approval date: November 12, 2024). The study was registered in the UMIN Clinical Trials Registry (UMIN000056499).

## Informed Consent Statement

Informed consent was obtained from all participants.

## Data Availability Statement

The data supporting the findings of this study are available upon request from the corresponding author. The data are not publicly available because they contain information that can compromise the privacy of the research participants.

## Acknowledgments

We would like to express our sincere gratitude to all the participants for their willingness to participate in this study. We gratefully acknowledge Mr. Tomohiko Mizuguchi at Maxell Ltd. for advice and suggestions regarding the analysis.

## References

1. United Nations. World Social Report 2023: Leaving No One Behind In An Ageing World, United Nations: New York, USA, 2023; pp. 17–75.

2. Miller, E.A.; Rosenheck, R.A.; Schneider, L.S. Caregiver burden, health utilities, and institutional service costs among community-dwelling patients with Alzheimer disease. Alzheimer Dis Assoc Disord 2010, 24, 380–389. doi: 10.1097/WAD.0b013e3181eb2f2e.

3. Cawthon, P.M.; Fox, K.M.; Gandra, S.R.; Delmonico, M.J.; Chiou, C.F.; Anthony, M.S.; Sewall, A.; Goodpaster, B.; Satterfield, S.; Cummings, S.R.; Harris, T.B. Health, Aging and Body Composition Study. Do muscle mass, muscle density, strength, and physical function similarly influence risk of hospitalization in older adults?. J Am Geriatr Soc 2009, 57, 1411–1419. doi: 10.1111/j.1532-5415.2009.02366.x.

4. World Health Organization. Decade of Healthy Ageing: Baseline Report, World Health Organization: Geneva, Switzerland, 2020; pp. 1–27.

5. Eckstrom, E.; Neukam, S.; Kalin, L.; Wright, J. Physical Activity and Healthy Aging. Clin Geriatr Med 2020, 36, 671–683. doi: 10.1016/j.cger.2020.06.009.

6. Kilbreath, S.L.; Heard, R.C. Frequency of hand use in healthy older persons. Aust J Physiother 2005, 51, 119–122. doi: 10.1016/s0004-9514(05)70040-4.

7. Incel, N.A.; Sezgin, M.; As, I.; Cimen, O.B.; Sahin, G. The geriatric hand: correlation of hand-muscle function and activity restriction in elderly. Int J Rehabil Res 2009, 32, 213–218. doi: 10.1097/MRR.0b013e3283298226.

8. Lin, C.H.; Chou, L.W.; Wei, S.H.; Lieu, F.K.; Chiang, S.L.; Sung, W.H. Influence of aging on bimanual coordination control. Exp Gerontol 2014, 53, 40–47. doi: 10.1016/j.exger.2014.02.005.

9. Murman, D.L. The Impact of Age on Cognition. Semin Hear 2015, 36, 111–121. doi: 10.1055/s-0035-1555115.

10. Rattanawan, P. Correlations between Hand Dexterity and Bimanual Coordination on the Activities of Daily Living in Older Adults with Mild Cognitive Impairment. Dement Geriatr Cogn Dis Extra 2022, 12, 24–32. doi: 10.1159/000521644.

11. Krehbiel, L.M.; Kang, N.; Cauraugh, J.H. Age-related differences in bimanual movements: A systematic review and meta-analysis. Exp Gerontol 2017, 98, 199–206. doi: 10.1016/j.exger.2017.09.001.

12. Kang, N.; Ko, D.K.; Cauraugh, J.H. Bimanual motor impairments in older adults: an updated systematic review and meta-analysis. EXCLI J 2022, 21, 1068–1083. doi: 10.17179/excli2022-5236.

13. Lee, T.D.; Wishart, L.R.; Murdoch, J.E. Aging, Attention, and Bimanual Coordination. Can J Aging/Rev Can Vieillissement 2002, 21, 549–557. doi:10.1017/S0714980800002087.

14. Rudisch, J.; Müller, K.; Kutz, D.F.; Brich L.; Sleimen-Malkoun R.; Voelcker-Rehage C. How Age, Cognitive Function and Gender Affect Bimanual Force Control. Front Physiol 2020, 11, 245. doi: 10.3389/fphys.2020.00245.

15. Chung, E.; Lee, S.H.; Lee, H.J.; Kim, Y.H. Comparative study of young-old and old-old people using functional evaluation, gait characteristics, and cardiopulmonary metabolic energy consumption. BMC Geriatr 2023, 23, 400. doi: 10.1186/s12877-023-04088-6.

16. Hayashida, I.; Tanimoto, Y.; Takahashi, Y.; Kusabiraki, T.; Tamaki, J. Correlation between muscle strength and muscle mass, and their association with walking speed, in community-dwelling elderly Japanese individuals. PLoS One 2014, 9, e111810. doi: 10.1371/journal.pone.0111810.

17. Kim, J.; Cha, E. Predictors of Cognitive Function in Community-Dwelling Older Adults by Age Group: Based on the 2017 National Survey of Older Korean Adults. Int J Environ Res Public Health 2021, 18, 9600. doi: 10.3390/ijerph18189600.

18. Blais, M.; Martin, E.; Albaret, J.M.; Tallet, J. Preservation of perceptual integration improves temporal stability of bimanual coordination in the elderly: an evidence of age-related brain plasticity. Behav Brain Res 2014, 275, 34–42. doi: 10.1016/j.bbr.2014.08.043.

19. Enokizono, T.; Ohto, T.; Tanaka, M.; Maruo, K.; Sano, Y.; Kandori, A.; Takada, H. Quantitative assessment of fine motor skills in children using magnetic sensors. Brain Dev 2020, 42, 421–430. doi: 10.1016/j.braindev.2020.03.004.

20. Tanaka, M.; Tsubouchi, M.; Kayashita, J.; Mizukami, K. [Factors associated with oral frailty among community-dwelling older people -A comparison between those <75 and ≥75 years old]. Nihon Ronen Igakkai Zasshi 2024, 61, 68–79. doi: 10.3143/geriatrics.61.68.

21. Suzumura S., Osawa A, Kanada Y., Keisuke M., Takano E., Sugioka J., Natsumi M., Nagahama T., Shiramoto K., Kuno K., Kizuka S., Satoh K., Sakurai H., Sano Y., Mizuguchi T., Kandori A, Kondo I. Finger tapping test for assessing the risk of mild cognitive impairment. Hong Kong J Occup Ther 2022, 35, 137–145. doi: 10.1177/15691861221109872.

22. Sugioka, J.; Suzumura, S.; Kuno, K.; Kizuka, S.; Sakurai, H.; Kanada, Y.; Mizuguchi, T.; Kondo, I. Relationship between finger movement characteristics and brain voxel-based morphometry. PLoS One 2022, 17, e0269351. doi: 10.1371/journal.pone.0269351.

23. Sano, Y.; Kandori, A.; Shima, K.; Tamura, Y.; Takagi, H.; Tsuji, T.; Noda, M.; Higashikawa, F.; Yokoe, M.; Sakoda, S. Reliability of Finger Tapping Test Used in Diagnosis of Movement Disorders. 2011 5th International Conference on Bioinformatics and Biomedical Engineering, Wuhan, China, 10-12 May 2011.

24. Suzumura, S.; Kanada, Y.; Osawa, A.; Sugioka, J.; Maeda, N.; Nagahama, T.; Shiramoto, K.; Kuno, K.; Kizuka, S.; Sano, Y.; Mizuguchi, T.; Kandori, A.; Kondo, I. Assessment of finger motor function that reflects the severity of cognitive function. Fujita Med J 2021, 7, 122–129. doi: 10.20407/fmj.2020-013.

25. Hu, X.; Newell, K.M. Aging, visual information, and adaptation to task asymmetry in bimanual force coordination. J Appl Physiol (1985) 2011, 111, 1671–1680. doi: 10.1152/japplphysiol.00760.2011.

26. Gerloff, C.; Andres, F.G. Bimanual coordination and interhemispheric interaction. Acta Psychol (Amst) 2002, 110, 161–186. doi: 10.1016/s0001-6918(02)00032-x.

27. Fling, B.W.; Seidler, R.D. Fundamental differences in callosal structure, neurophysiologic function, and bimanual control in young and older adults. Cereb Cortex 2012, 22, 2643–2652. doi: 10.1093/cercor/bhr349.

28. Fujiyama, H.; Van Soom, J.; Rens, G.; Gooijers, J.; Leunissen, I.; Levin, O.; Swinnen, S.P. Age-Related Changes in Frontal Network Structural and Functional Connectivity in Relation to Bimanual Movement Control. J Neurosci 2016, 36, 1808–1822. doi: 10.1523/JNEUROSCI.3355-15.2016.

29. Li, S.C.; Lindenberger, U.; Sikström, S. Aging cognition: from neuromodulation to representation. Trends Cogn Sci 2001, 5, 479–486. doi: 10.1016/s1364-6613(00)01769-1.

30. Verdú, E.; Ceballos, D.; Vilches, J.J.; Navarro, X. Influence of aging on peripheral nerve function and regeneration. J Peripher Nerv Syst 2000, 5, 191–208. doi: 10.1046/j.1529-8027.2000.00026.x.

31. Banerjee, A.; Jirsa, V.K. How do neural connectivity and time delays influence bimanual coordination?. Biol Cybern 2007, 96, 265–278. doi: 10.1007/s00422-006-0114-4.

32. Wilson, T.W.; Kurz, M.J.; Arpin, D.J. Functional specialization within the supplementary motor area: a fNIRS study of bimanual coordination. Neuroimage 2014, 85, 445–450. doi: 10.1016/j.neuroimage.2013.04.112.

33. Green, P.E.; Ridding, M.C.; Hill, K.D.; Semmler, J.G.; Drummond, P.D.; Vallence, A.M. Supplementary motor area-primary motor cortex facilitation in younger but not older adults. Neurobiol Aging 2018, 64, 85–91. doi: 10.1016/j.neurobiolaging.2017.12.016.

34. Cansino, S.; Hernández-Ramos, E.; Estrada-Manilla, C.; Torres-Trejo, F.; Martínez-Galindo, J.G.; Ayala-Hernández, M.; Gómez-Fernández, T.; Osorio, D.; Cedillo-Tinoco, M.; Garcés-Flores, L.; Beltrán-Palacios, K.; García-Lázaro, H.G.; García-Gutiérrez, F.; Cadena-Arenas, Y.; Fernández-Apan, L.; Bärtschi, A.; Rodríguez-Ortiz, M.D. Decline in verbal and visuospatial working memory across adult lifespan. Age (Dordr) 2013, 35, 2283–2302. doi: 10.1007/s11357-013-9531-1.

35. Ingemanson, M.L.; Rowe, J.B.; Chan, V.; Wolbrecht, E.T.; Cramer, S.C.; Reinkensmeyer, D.J. Use of a robotic device to measure age-related decline in finger proprioception. Exp Brain Res 2016, 234, 83–93. doi: 10.1007/s00221-015-4440-4.

36. Mickevičienė, D.; Skurvydas, A.; Karanauskienė, D. Is intraindividual variability different between unimanual and bimanual speed-accuracy movements?. Percept Mot Skills 2015, 120, 125–138. doi: 10.2466/25.PMS.120v14×3.

37. Swinnen, S.P. Intermanual coordination: from behavioural principles to neural-network interactions. Nat Rev Neurosci 2002, 3, 348–359. doi: 10.1038/nrn807.

38. Johansen-Berg, H.; Della-Maggiore, V.; Behrens, T.E.; Smith, S.M.; Paus, T. Integrity of white matter in the corpus callosum correlates with bimanual co-ordination skills. Neuroimage 2007, 36, 16–21. doi: 10.1016/j.neuroimage.2007.03.041.

39. Fling, B.W.; Walsh, C.M.; Bangert, A.S.; Reuter-Lorenz, P.A.; Welsh, R.C.; Seidler, R.D. Differential callosal contributions to bimanual control in young and older adults. J Cogn Neurosci 2011, 23, 2171–2185. doi: 10.1162/jocn.2010.21600.

40. Kiyama, S.; Kunimi, M.; Iidaka, T.; Nakai, T. Distant functional connectivity for bimanual finger coordination declines with aging: an fMRI and SEM exploration. Front Hum Neurosci 2014, 8, 251. doi: 10.3389/fnhum.2014.00251.

41. Nicolay, C.W.; Walker, A.L. Grip strength and endurance: Influences of anthropometric variation, hand dominance, and gender. Int J Ind Ergon 2005, 35, 605–618. doi: 10.1016/j.ergon.2005.01.007.

42. Józsa, L.; Demel, Z.; Vándor, E.; Réffy, A.; Szilágyi, I. Spezifische Faserzusammensetzung der menschlichen Handund Armmuskeln [Specific fibre compositon of human hand and arm muscles]. Handchirurgie 1978, 10, 153–157.

43. Bütefisch, C.M.; Khurana, V.; Kopylev, L.; Cohen, L.G. Enhancing encoding of a motor memory in the primary motor cortex by cortical stimulation. J Neurophysiol 2004, 91, 2110–2116. doi: 10.1152/jn.01038.2003.

44. Hodges, L.; Adams, J. Grip Strength and Dexterity: A Study of Variance between Right- and Left-Handed Healthy Individuals. The British Journal of Hand Therapy 2007, 12, 15–21. doi:10.1177/175899830701200102.

45. Weller, M.P.; Latimer-Sayer, D.T. Increasing right hand dominance with age on a motor skill task. Psychol Med 1985, 15, 867–872. doi: 10.1017/s0033291700005109.

46. Zhou, G.; Chen, Y.; Wang, X.; Wei, H.; Huang, Q.; Li, L. The correlations between kinematic profiles and cerebral hemodynamics suggest changes of motor coordination in single and bilateral finger movement. Front Hum Neurosci 2022, 16: 957364. doi: 10.3389/fnhum.2022.957364.

47. Li, S.; Danion, F.; Latash, M.L.; Li, Z.; Zatsiorsky, V.M. Finger Coordination and Bilateral Deficit during Two-Hand Force Production Tasks Performed by Right-Handed Subjects. J Appl Biomech 2000, 16, 379–391. doi:10.1123/jab.16.4.379.

48. Tomita, Y.; Tanaka, S.; Takahashi, S.; Takeuchi, N. Detecting cognitive decline in community-dwelling older adults using simple cognitive and motor performance tests. Geriatr Gerontol Int 2020, 20, 212–217. doi: 10.1111/ggi.13863.

